# Rapid Diffusion, Persistent Deserts: A National Longitudinal Study of Geographic Access to Hospitals with AI Deployment, 2022–2024

**DOI:** 10.64898/2026.04.28.26351957

**Authors:** Aaron Johnson, David Gefen, Teresa Harrison

## Abstract

Measured hospital AI deployment expanded between 2022 and 2024, but did the geography of access converge, and did persistent access deserts overlap with greater health burden? Using two waves of the American Hospital Association (AHA) Annual Survey linked to 2020 Census block-group populations, we estimate contiguous-U.S. coverage for 329.3 million residents and full-frame transition profiles for 334.7 million residents. Measured AI-enabled status is identified using five binary workforce/workflow AI-use items in 2022 and fourteen ordinal clinical and operational AI implementation items in 2024 (primary 2024 threshold: expanding or fully integrated).

The share of hospitals reporting active AI deployment rose from 18.3% to 28.6%. Contiguous-U.S. coverage within a 30-minute drive increased from 67.0% to 76.1%, yet spatial inequality grew: the population-weighted Gini coefficient of access distances rose from 0.739 to 0.767. In the full transition frame, 45.1 million people newly crossed the 30-minute threshold, while 67.9 million remained outside it in both waves: 37.2 million experienced no travel-time improvement and 30.7 million improved but still did not cross the threshold.

These findings reveal a diffusion paradox: measured expansion coexists with persistent and, by some measures, rising inequality in who benefits. The communities left behind are not randomly distributed; they are more rural, lower-income, higher-poverty, older, more uninsured, and carry higher baseline premature-mortality burden than persistently served communities. A pre-diffusion Years of Potential Life Lost (YPLL) check showed that the mortality-burden gradient was already present before the 2022–2024 diffusion window, supporting a burden-overlap interpretation rather than causal mortality evidence.

Significance Statement
Using two AHA survey waves (2022–2024), we estimate contiguous-U.S. 30-minute coverage for 329.3 million residents and full-frame transition profiles for 334.7 million residents. Coverage rose from 67.0% to 76.1%, yet spatial inequality increased (Gini 0.739 to 0.767; bootstrap *p <* 0.05). In the full transition frame, 67.9 million people remained outside 30-minute access in both waves, including 37.2 million with no travel-time improvement. Persistently excluded communities also carried higher baseline premature mortality, and a pre-diffusion check showed that this gradient was already present before the 2022–2024 diffusion window, making the access pattern a health-equity concern rather than only a geography-of-technology finding.

## 1 Introduction

When a potentially valuable hospital capability diffuses rapidly, does access become more equal or less equal? Hospital AI encompasses both *operational* tools—staffing optimization, scheduling, demand forecasting, and routine-task automation— and *clinical* applications such as diagnostic, decision-support, and population-health AI.^1,2^ We define a hospital as AI-enabled if it reports active deployment (expanding or fully integrated) on any of 14 ordinal implementation items spanning six operational and eight clinical domains in the 2024 AHA Annual Survey, or reports any of five binary AI/ML workforce items in 2022 (full crosswalk in Methods and SI Section S1).

This study separates *adoption prevalence*—the share of hospitals reporting AI—from *population coverage*—the share of people who can reach an AI-deploying hospital within a policy-relevant time threshold. ^3^ Early adoption tends to cluster in metropolitan hubs, so modest adoption can still translate into high coverage while leaving large rural regions effectively unserved. ^4^ This mismatch means technology diffusion can raise national coverage while *increasing* spatial concentration, leaving persistent “access deserts” even as average distance falls. Importantly, general hospital geography did not change materially between our two survey waves (all-hospital access Gini ≈ 0.831 in both years), so the patterns we document reflect where AI is layered onto an otherwise stable hospital landscape, not general hospital closure or consolidation.

Because causal evidence on AI’s clinical effects remains limited, the equity question is not whether AI deployment can yet be credited with better outcomes, but whether access to an expanding capability is being layered onto communities already carrying socioeconomic and health burdens. ^5,6^Premature mortality sharpens this question. We use baseline Years of Potential Life Lost (YPLL) as descriptive health-burden context: do persistent AI access deserts overlap with places already carrying elevated premature mortality, and does that mortality gradient predate the 2022–2024 diffusion window? This distinction matters because the policy relevance of diffusion depends not only on how many people gain access, but on whether those left outside timely access are communities already facing greater health and socioeconomic burden.

We build a two-wave longitudinal design using U.S. hospital and county data for 2022 and 2024, quantifying diffusion at both the county and Census block-group level. We estimate national expansion, inequality (Lorenz/Gini), and upper-tail remoteness (P90, the distance above which the worst-served 10% of people live), and we track how many people cross a policy-relevant 30-minute access threshold. The central contribution is a national two-wave portrait of AI diffusion, spatial inequality, and health-burden overlap that explicitly separates “broad improvement” from “threshold crossing” and characterizes the communities that gained access versus those persistently excluded.

The framework draws on Diffusion of Innovation theory, ^7^ which predicts non-uniform spread and implies that growth and inequality can move in opposite directions—a diffusion paradox. We use nearest-facility and threshold-coverage measures because they are interpretable as emergency-access benchmarks and align with diffusion questions about where the first AI-enabled facility appears; supply-sensitive alternatives (E2SFCA) are reported as benchmarks in the Limitations and SI. ^8–11^

### Research Questions

#### RQ1 Diffusion and inequality

How did geographic access to AI-deploying hospitals change between 2022 and 2024, and did spatial inequality narrow or widen?

#### RQ2 Who gained and who remained excluded

Who newly gained timely access, who remained persistently outside 30-minute access, and how do those communities differ in socioeconomic and health characteristics?

#### RQ3 Health-burden overlap

Do persistent AI access deserts overlap with higher-YPLL communities, and did that gradient predate the 2022–2024 AI diffusion window?

We treat YPLL as descriptive health-burden context, not as a causal outcome of AI deployment.

## 2 Results

### A. Reported Deployment Expanded, but Inequality Worsened

#### Longitudinal Adoption, Coverage, and Inequality

Table 1 shows measured AI expansion layered onto a stable hospital geography. In the county-panel frame, AI-enabled hospitals increased from 1,119 to 1,743 (+56% relative), and AI 30-minute coverage increased from 66.2% to 75.2%. However, AI access inequality rose (Gini 0.739 to 0.767), while baseline hospital geography was effectively unchanged (all-hospital Gini ≈ 0.831 in both years). Notably, adoption prevalence and coverage diverge: only 28.6% of hospitals report active AI deployment, yet 75.2% of the population are within 30 minutes, underscoring why diffusion should be evaluated in *people reached* rather than hospitals counted. ^12^

**Table 1.** Longitudinal diffusion and inequality summary (county panel, 2022–2024; full analysis frame, population base ≈334.7 million).

| Metric | 2022 | 2024 | Change |
| --- | --- | --- | --- |
| AI-enabled hospitals (n) | 1,119 | 1,743 | +624 |
| AI-enabled hospitals (%) | 18.3 | 28.6 | +10.3 pp |
| AI population covered (%) | 66.2 | 75.2 | +9.0 pp |
| AI access Gini | 0.739 | 0.767 | +0.028 |
| All-hospital access Gini | 0.832 | 0.831 | $\approx 0$ |

*Notes:* County-panel summary. Coverage in this panel uses the full analysis frame (population base $\approx 334.7$ million). “All-hospital access Gini” is the population-weighted Gini coefficient of distances to the nearest hospital of *any* type (regardless of technology), serving as a baseline measure of underlying hospital geography. Contiguous-U.S. block-group coverage is reported separately in Table 3.

The AI access Gini *increased* (0.739 to 0.767) even as the all-hospital access Gini was essentially unchanged, indicating that the inequality increase reflects where AI was layered onto the hospital system rather than broader hospital geography. Census-division heterogeneity reinforces this pattern, with AI adoption gains ranging from +5.3 pp (West South Central) to near-zero (Middle Atlantic; Table S2 in SI). Alternative distributional metrics yielded the same qualitative conclusion: the AI Atkinson index rose from 0.477 to 0.526 and the AI Theil index rose from 1.43 to 1.73, while all-hospital metrics were essentially stable (Table S9 in SI).

#### Distance-Gap Dynamics: Compression with Persistent Concentration

The population-weighted P90–P10 distance gap for AI fell from 49.8 to 33.2 miles (Table 2), indicating meaningful upper-tail improvement—yet the Gini rose. This compression-with-concentration pattern reflects metropolitan-focused adoption that fattens the near-zero distance mass while the far tail shrinks but remains large (stratification by USDA Rural-Urban Continuum Code [RUCC] group in Figure 3 confirms the metro–rural divergence). By contrast, the all-hospital P90–P10 gap was essentially unchanged, confirming that these dynamics are specific to AI diffusion.

**Table 2.** Population-weighted P10/P90 distance and gap change (miles).

| Access Type | P10 (2022) | P10 (2024) | P90 (2022) | P90 (2024) | Gap Change |
| --- | --- | --- | --- | --- | --- |
| All hospitals | 0.84 | 0.85 | 10.55 | 10.57 | +0.02 |
| AI | 1.80 | 1.42 | 51.61 | 34.66 | -16.57 |

*Notes:* P10 is the travel distance below which the best-served 10% of people live, and P90 is the distance above which the worst-served 10% live. A larger P90–P10 gap means the worst-served communities are much farther from AI-enabled hospitals than the best-served communities.

### B. Threshold Crossing, Persistent Exclusion, and Who Was Left Behind

Contiguous-U.S. block-group mapping (236,879 block groups; population 329.3 million) shows similar gains to the county-panel headline rates. AI hospitals rose from 1,119 to 1,743 nationally, and the contiguous-frame share within 30 minutes rose from 67.0% to 76.1% (+29.9 million people).

The threshold-versus-continuous contrast is central: 143.5 million people improved in drive time, but only 44.7 million crossed into ≤30-minute access. In 2024, approximately 79 million people remained outside 30 minutes (mean drive time 73.0 min).

Because maps, transition profiles, and robustness checks use different fixed analytic frames, three population counts should be read separately: 329.3 million for contiguous-U.S. maps, 334.7 million for full-frame transition profiles, and 67.9 million for the outside-both-waves transition group after excluding likely instrument-reclassified lost access.

Figure 2 separates threshold crossing from continuous improvement. The left panel identifies where block groups newly crossed into ≤30-minute access, while the right panel shows net reductions in proxy drive time, including areas that improved but did not cross the threshold.

**Fig. 1.**
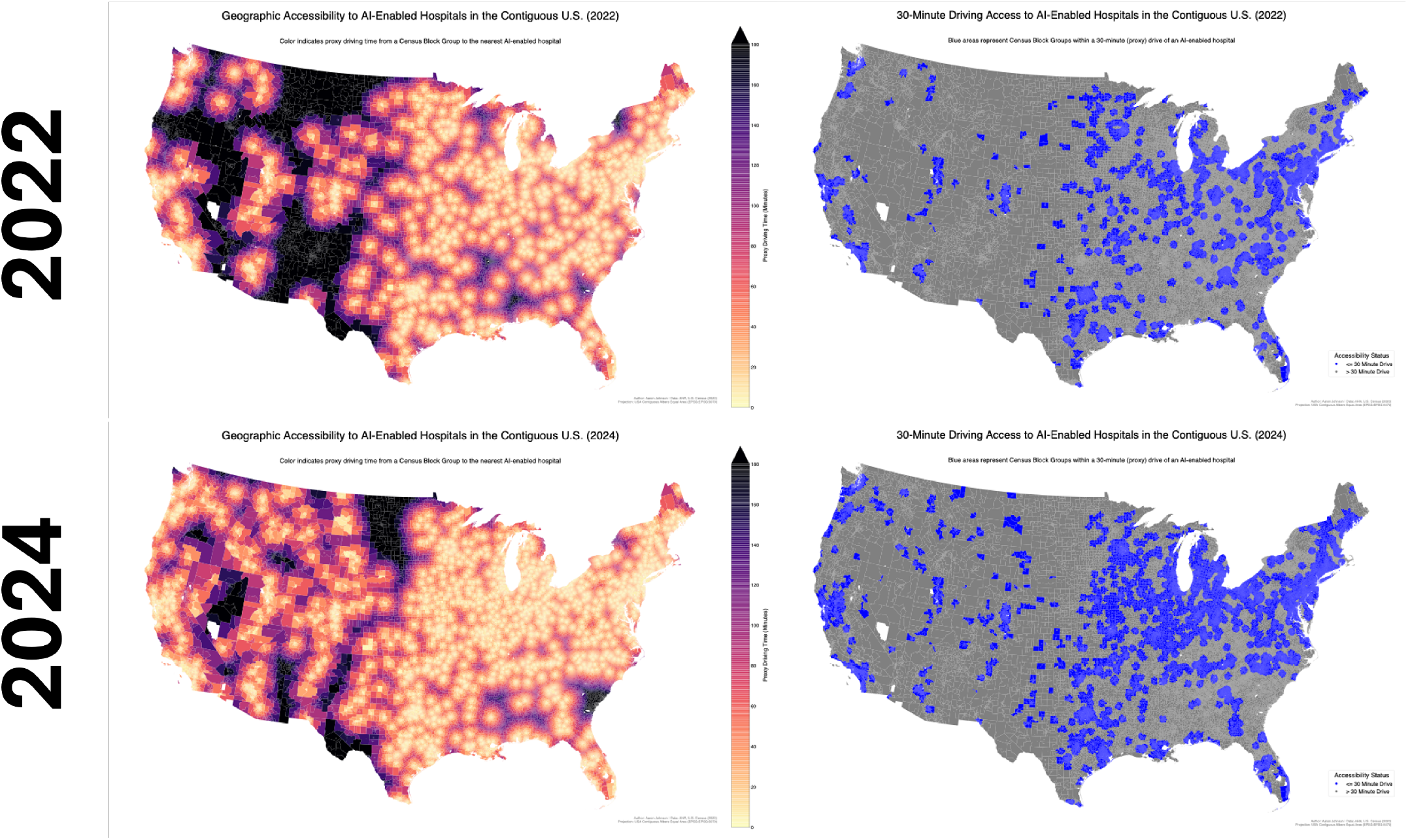
National diffusion in AI-enabled hospital access from 2022 to 2024 (contiguous-U.S. block-group frame; 236,879 block groups; 329.3 million people). The top row shows 2022 and the bottom row shows 2024 for contiguous-U.S. census block groups. Left panels report continuous proxy driving time to the nearest AI-enabled hospital (haversine distance converted to minutes using a 40 mph speed assumption and 1.3 circuity factor; see Section 4.2 for validation); darker tones indicate longer travel time. Right panels show binary accessibility within 30 minutes. Coverage expands substantially between waves, especially across the Midwest, Great Lakes, Appalachia, and parts of the South, while persistent access deserts remain in the interior West and northern Plains.

**Fig. 2.**
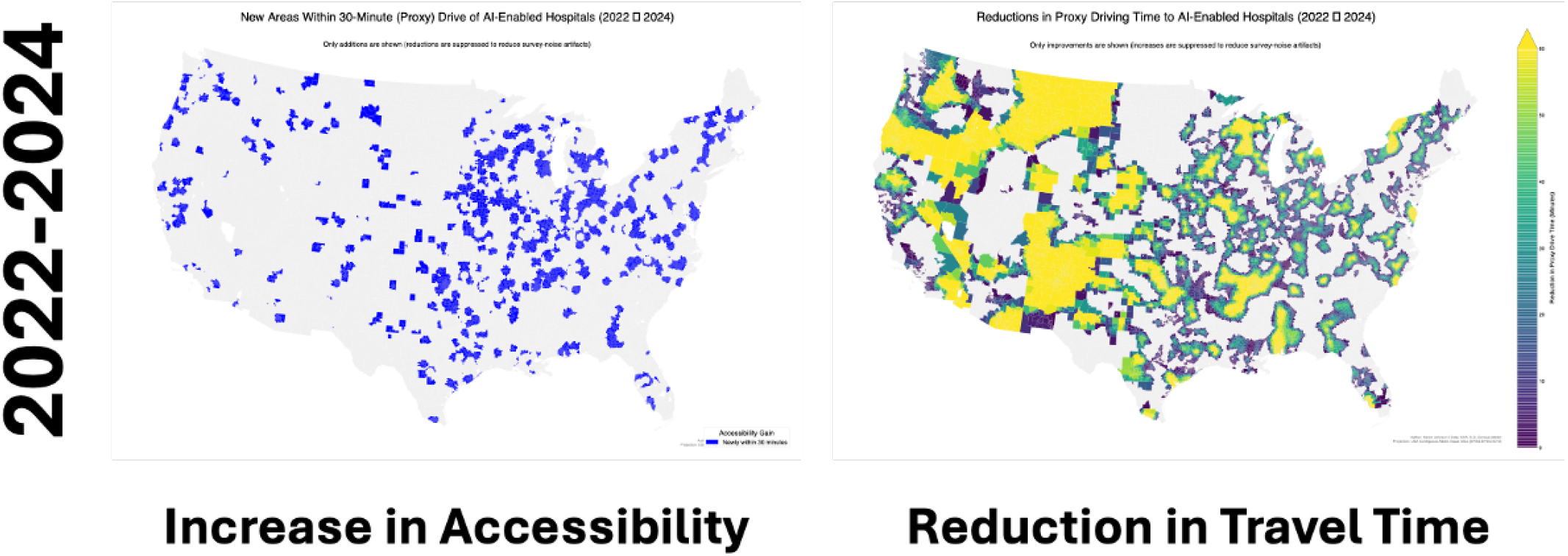
Block-group access gains from 2022 to 2024: threshold crossings versus continuous time reduction. Left panel: census block groups in the contiguous U.S. that became newly within a 30-minute proxy drive to an AI-enabled hospital. Right panel: net reduction in proxy driving time to the nearest AI-enabled hospital (minutes), mapped for improvements only. The two panels show that diffusion produced both discrete policy-relevant gains (newly within 30 minutes) and broader continuous accessibility improvements across many additional areas that remain beyond the threshold.

**Fig. 3.**
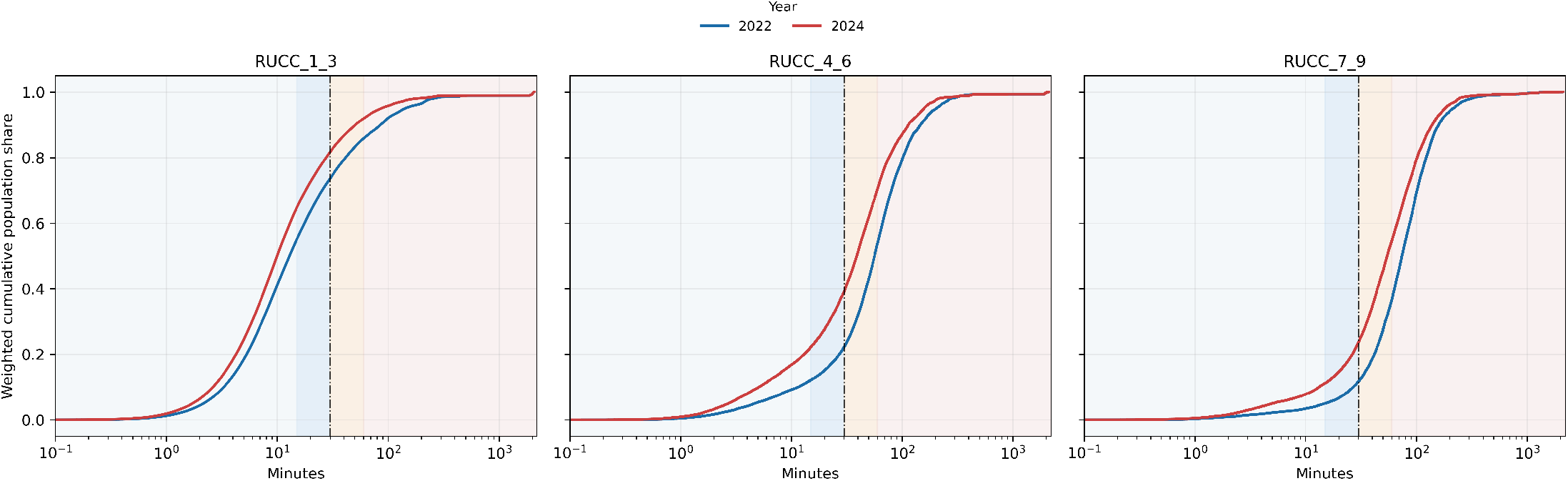
Travel-time CDF by RUCC group, 2022 vs. 2024. Population-weighted cumulative distribution of proxy travel time to the nearest AI-deploying hospital, stratified by metro (RUCC 1–3), more-urbanized nonmetro (RUCC 4–6), and less-urbanized nonmetro (RUCC 7–9). These are rurality-size bins rather than ERS metro-adjacency bins. The metro curve shifts dramatically left between waves, while the less-urbanized nonmetro curve moves only slightly. This stratification visualizes the rural–urban divergence underlying the aggregate Gini increase.

To move beyond aggregate counts, Table 4 profiles the populations in each access-transition group by county-linked demographic and socioeconomic context. An additional 10,861 block groups (14.9 million people) appeared within 30 minutes in 2022 but fell outside the threshold in 2024. Because the AHA Annual Survey transitioned from binary to ordinal AI items between survey waves, we classify these block groups separately as an apparent “lost-access” group rather than combining possible instrument reclassification with stable access deserts.

**Table 3.** Contiguous-U.S. block-group access summary (2022 vs 2024; 236,879 block groups; population 329.3 million).

| Metric | 2022 | 2024 |
| --- | --- | --- |
| Block groups (n) | 236,879 | 236,879 |
| AI-enabled hospitals (n) | 1,119 | 1,743 |
| Population within 30 min (millions) | 220.7 (67.0%) | 250.6 (76.1%) |
| Mean proxy drive time (min) | 36.4 | 25.1 |
| Median proxy drive time (min) | 15.3 | 11.3 |
| P90 proxy drive time (min) | 94.8 | 62.9 |

*Notes:* AI-enabled hospital counts are from the full national analysis frame (Table 1); coverage rates and drive-time statistics are computed on the contiguous-U.S. block-group frame. *Change diagnostics (2022–2024):* newly within 30 minutes = 32,965 block groups (44.7 million people); any improvement = 104,493 block groups (143.5 million people); improved but did not newly cross into $\leq 30$ -minute access = 71,528 block groups (98.8 million people), which includes both block groups already within 30 minutes that experienced further drive-time reductions and the 24,745 block groups (30.7 million people) that improved but remained outside 30 minutes (Table 4); still outside 30 minutes in 2024 = 61,639 block groups (approximately 79 million people).

**Table 4.** Demographic and socioeconomic profile of access-transition groups, 2022–2024 (full analysis frame; 240,938 block groups; 334.7 million people; 10,861 lost-access block groups excluded from “outside, no improvement” column). The “improved, still outside” and “outside, no improvement” groups together constitute the 67.9 million people outside 30-minute access in both waves. Note: the four column populations (206.8 + 45.1 + 30.7 + 37.2 = 319.8 million) plus the excluded 14.9 million lost-access group sum to the full-frame total; the approximately 79 million outside 30 minutes in 2024 reported in Table 3 is a contiguous-frame cross-section and is not directly additive with these longitudinal transition groups.

| Characteristic | Persistently within<br>30 min (142,937 BGs)<br>206.8 million | Newly within<br>30 min (33,259 BGs)<br>45.1 million | Improved, still<br>outside (24,745 BGs)<br>30.7 million | Outside, no<br>improvement (29,136 BGs)<br>37.2 million |
| --- | --- | --- | --- | --- |
| *Rurality (RUCC, population-weighted %)* |  |  |  |  |
| Metro (RUCC 1–3) | 97.0 | 79.4 | 52.6 | 58.2 |
| Nonmetro, more urban (RUCC 4–6) | 2.3 | 14.7 | 26.5 | 24.8 |
| Nonmetro, less urban (RUCC 7–9) | 0.6 | 6.0 | 20.9 | 17.0 |
| *Socioeconomic context (county-linked, population-weighted mean)* |  |  |  |  |
| Median household income (\$) | 84,046 | 70,656 | 65,018 | 61,386<sup>†</sup> |
| Poverty rate (%) | 12.7 | 14.1 | 15.8 | 15.7<sup>†</sup> |
| Uninsured rate (%) | 9.7 | 10.8 | 11.1 | 11.8<sup>†</sup> |
| Social & economic factors index | 11.0 | 13.1 | 13.8 | 14.5<sup>†</sup> |
| *Health and demographics (county-linked, population-weighted mean)* |  |  |  |  |
| Health behaviors index | 44.3 | 39.1 | 35.1 | 36.1<sup>†</sup> |
| % population aged 65+ | 14.5 | 16.8 | 18.1 | 18.7<sup>†</sup> |
| Baseline county YPLL/100k (2019; descriptive context) | 6,375 | 7,456 | 8,219 | 8,935<sup>†</sup> |
| *Race/ethnicity (population-weighted %)* |  |  |  |  |
| Non-Hispanic White | 55.2 | 67.4 | 72.1 | 77.5<sup>†</sup> |
| Non-Hispanic Black | 14.0 | 9.9 | 8.8 | 11.5<sup>†</sup> |
| Hispanic/Latino | 20.7 | 16.2 | 12.8 | 7.4<sup>†</sup> |
| *Access metrics (population-weighted)* |  |  |  |  |
| Mean proxy drive time 2024 (min) | 9.3 | 13.4 | 65.9 | 251.8 |
| Median proxy drive time 2024 (IQR, min) | 7.6 (4.1–12.7) | 12.3 (6.0–20.3) | 50.7 (38.1–76.0) | 64.1 (42.6–121.1) |
| Mean proxy drive time 2022 (min) | 10.6 | 97.4 | 109.3 | 234.3 |
| Median proxy drive time 2022 (IQR, min) | 8.8 (4.9–15.0) | 71.6 (44.9–113.2) | 86.2 (58.6–131.1) | 52.3 (38.5–85.3) |
| Mean $\Delta$ drive time (min) | –1.3 | –84.0 | –43.4 | +17.5<sup>‡</sup> |

Notes: BGs = census block groups. Block groups classified by AI access transition between 2022 and 2024 using the 30-minute proxy-drive-time threshold. “Outside, no improvement” includes only block groups that were outside 30 minutes in both waves and did not experience any travel-time reduction; “improved, still outside” block groups were also outside in both waves but experienced reduced travel time. Together these two groups constitute all 67.9 million people outside 30 minutes in both waves. An additional 10,861 block groups (14.9 million people) that appeared within 30 minutes in 2022 but fell outside in 2024 are excluded from the “outside, no improvement” column because the AHA Annual Survey transitioned from binary to ordinal AI items between survey waves; the data cannot distinguish instrument reclassification from genuine service withdrawal for this apparent “lost-access” group. County-linked characteristics assigned via county FIPS. RUCC = USDA Rural-Urban Continuum Codes. RUCC 4–6 and 7–9 are rurality-size bins rather than ERS metro-adjacency bins. Social & economic factors index and health behaviors index from County Health Rankings (higher values = more disadvantaged for social/economic; lower = less favorable health behaviors).
<sup>†</sup> County-linked demographic values for the outside/no-improvement column are adjusted estimates net of the excluded lost-access subgroup; drive-time metrics are computed directly from block-group data. <sup>‡</sup> The high mean drive times for the outside/no-improvement group ( $\approx 234$ –252 minutes) are not inconsistent with the contiguous-frame P90 of $\approx 63$ minutes in 2024 (Table 3): the persistently-outside group represents the extreme tail ( $\approx 11\%$ of the population), well beyond the 90th percentile, and includes extremely remote block groups in the full national frame with drive times exceeding 500 minutes that raise the population-weighted mean substantially above the median reported in the table. The positive mean $\Delta$ for the outside/no-improvement group is sensitive to the AHA instrument change: hospitals that qualified as AI-enabled under the 2022 binary items but fell below the “active deployment” threshold (stage $\geq 3$ , expanding or fully integrated) on the 2024 ordinal items ceased to count as AI-enabled, pushing the nearest qualifying hospital farther away for some already-remote block groups. This is consistent with the instrument-change concern described in the Limitations; the increase is reduced to +8.3 minutes under the broader “any activity” threshold and is not observed under the operational-only sensitivity restriction.

In total, 67.9 million people remained outside 30-minute access in both waves. Of these, 30.7 million experienced travel-time improvement but did not cross the threshold, and 37.2 million experienced no improvement (or worsened). The no-improvement group lives in counties with markedly lower median household income ($61,386 vs. $84,046), higher poverty (15.7% vs. 12.7%), higher uninsurance (11.8% vs. 9.7%), and substantially more nonmetropolitan geography (41.8% non-metro vs. 3.0%) compared to the persistently served. This gradient is broadly monotonic: on nearly every socioeconomic indicator, the four transition groups—persistently within, newly within, improved-but-outside, and persistently outside—form an ordered sequence from most to least advantaged (the sole exception is poverty rate, which is 15.8% for improved-but-outside and 15.7% for persistently outside). The 45.1 million who newly crossed the threshold occupy an intermediate position—substantially more rural than the persistently served (20.6% nonmetro vs. 3.0%), but less disadvantaged than those persistently excluded. The no-improvement group’s mean proxy drive time in 2024 was 251.8 minutes—over four hours—but the population-weighted median was 64.1 minutes (IQR 42.6– 121.1), highlighting a heavily right-skewed distribution in which extremely remote block groups inflate the mean. This underscores the severity of the access deficit for communities that diffusion has not reached.

### C. Health-Burden Overlap in Persistent Access Deserts

Table 4 also makes premature mortality a main descriptive equity-burden finding. Baseline county YPLL rises across the transition profile: 6,375 per 100,000 among persistently served communities, 7,456 among newly served communities, 8,219 among communities that improved but remained outside 30-minute access, and 8,935 among the outside/no-improvement group. A county-level check in SI Section S4 shows that the distance–YPLL gradient predates the 2022–2024 diffusion window: 2024 AI distance is associated with both 2024 YPLL and 2019 YPLL. These results indicate that AI access deserts overlap with pre-existing premature-mortality burden; they do not identify a causal effect of AI deployment on mortality.

### D. Sensitivity to AI Definition

Each sensitivity test addresses a specific alternative explanation for the main pattern.

**Threshold sensitivity** addresses the concern that the inequality finding depends on where the adoption bar is set. Moving from a broader *any activity* definition (exploring or above; analysis stage ≥ 1) to *active deployment* (expanding or fully integrated; stage ≥ 3) reduces measured prevalence (39.4% to 28.6%) and coverage (80.7% to 75.2%) while preserving high inequality (Gini 0.780 to 0.767), supporting the active-deployment threshold as a conservative main specification (Figure S1 in SI). A further restriction to fully integrated only (stage ≥ 4) halves the hospital count to 859 (14.1%) and drops coverage to 58.5%, yet the Gini remains above 0.73 (Table S4 in SI). Across all three thresholds, the core finding—measured expansion with persistent inequality—is unchanged.

**Access-threshold sensitivity** addresses the concern that some hospital AI capabilities are not emergency-sensitive and may be meaningfully reachable beyond 30 minutes. Repeating the transition analysis at 45 and 60 minutes increases full-frame 2024 coverage to 279.2 million and 295.4 million people, respectively, but 41.1 million and 27.2 million people still remain outside both waves after excluding apparent lost access (Table S5 in SI). Thus, longer catchments reduce the absolute desert population, as expected, without eliminating the core transition pattern.

**Construct-comparability anchor: operational-only items** addresses the concern that the 2024 instrument expansion (adding eight clinical AI items with no 2022 counterpart) mechanically inflates measured diffusion. The most direct cross-wave comparison restricts the 2024 flag to the six *operational* items only at the active-deployment threshold (stage ≥ 3, expanding or fully integrated), excluding all clinical AI columns (Table S3 in SI). Under this “common-item” restriction, 1,091 hospitals (18.0%) qualify and coverage drops to 63.6%—notably, operational-only 2024 prevalence (18.0%) is virtually identical to the 2022 headline (18.3%), indicating that the measured expansion is driven largely by the clinical AI items added in the 2024 instrument rather than common-item operational AI growth. Crucially, the Gini remains high at 0.747 (cf. 0.767 for the full 14-item flag), confirming that the inequality finding is robust even when the diffusion component is minimal. A full replication of the transition-group analysis under the operational-only restriction (Table S10 in SI) raises the outside/no-improvement plus lost-access group from 52.2 to 93.0 million (+78.3%). Relative to the main adjusted persistently-outside benchmark of 67.9 million, the operational-only estimate is 37% larger. This denominator distinction underscores that clinical AI items unique to 2024 substantially expand measured coverage while the core inequality pattern is preserved.

**Tight 2022 denominator sensitivity** addresses the possibility that the binary 2022 workforce/workflow fields are too permissive. In the geocoded spatial-analysis subset, requiring at least two affirmative 2022 AI items reduces 2022 AI hospitals from 1,118 to 878 and 30-minute coverage from 221.7 million to 207.2 million people, while the 2022-tight-to-2024-primary transition still leaves 70.5 million people outside both waves (Table S6 in SI). This check strengthens the measurement interpretation: a stricter 2022 denominator changes the level of measured expansion but does not remove the persistent access-desert result.

**Construct-comparability anchor: clinical-only items** addresses the mirror concern—that the inequality finding is driven by operational AI items that may reflect generic workflow automation rather than clinically meaningful capability. Restricting the 2024 flag to the eight *clinical* AI items only yields 1,557 hospitals (25.6%), coverage of 73.4%, and a Gini of 0.765— nearly unchanged from the full-definition estimate, confirming robustness to construct definition (Table S12 in SI).

## 3 Discussion

The main empirical message is a diffusion paradox: measured hospital AI deployment expanded between 2022 and 2024, tail remoteness improved, but full-distribution inequality did not converge. Persistent access deserts remain, concentrated in communities that are simultaneously more rural, lower-income, higher-poverty, older, and higher in baseline premature-mortality burden than persistently served communities (Table 4). At stake is not merely a geographic inconvenience: 67.9 million people remained outside 30-minute access in both waves, of whom 37.2 million experienced no travel-time improvement and 30.7 million improved but did not cross the threshold. Even the better-off subgroup lives in counties with median household income $19,000 below the persistently served, while the no-improvement group faces a $23,000 income gap, poverty rates 24% higher, uninsured rates 22% higher, and 40% higher baseline YPLL, with AI access exclusion layered onto pre-existing socioeconomic and health burden. The central contribution is empirical: a national two-wave portrait that explicitly separates broad improvement from threshold crossing, characterizes the communities that gained access versus those persistently excluded, and shows that persistent AI access deserts overlap with pre-existing premature-mortality burden.

The Gini increase (0.739 to 0.767) warrants careful interpretation. The P90–P10 gap fell substantially while the Gini rose—a compression-with-concentration pattern: metropolitan-focused adoption fattens the near-zero distance mass while leaving the far tail relatively unchanged. County-clustered bootstrap resampling confirms this increase is significant (change 0.028; 95% CI [0.016, 0.038]; Table S8 in SI), whereas the all-hospital Gini change is effectively zero, indicating that the rising inequality is specific to AI deployment geography. A supply-sensitive E2SFCA benchmark using staffed beds and a 30-minute Gaussian catchment yields the same qualitative pattern: median access declines from metropolitan to less-urbanized nonmetro areas, the less-urbanized nonmetro P10 remains zero in both waves, and geographic gradients are broadly concordant with the nearest-facility results (Section S11). Census-division heterogeneity (Table S2) confirms broad but uneven adoption growth: nearly all divisions gained adoption share, while the Middle Atlantic was essentially flat.

### 3.1 Who benefited, who was left behind, and why thresholds matter

Two facts jointly describe who gained: (1) 44.7 million people newly crossed the 30-minute threshold (contiguous-U.S. frame; 45.1 million in the full national frame of Table 4), and (2) another 98.8 million people experienced shorter travel times without newly crossing the threshold. That second group combines two substantively different populations: 30.7 million people who remained outside 30-minute access despite improvement, and approximately 68.1 million people who were already within 30 minutes and became even closer. A binary threshold alone misses both forms of progress, while continuous distance alone can hide policy-relevant discontinuities: for many communities, the difference between 35 and 25 minutes is a qualitatively different access state.

The transition-group profiling (Table 4) adds an equity dimension. Of the 67.9 million people outside 30-minute access in both waves, the 37.2 million who experienced no travel-time improvement are the most disadvantaged: disproportionately in nonmetropolitan counties (42% nonmetro vs. 3% for the persistently served), with higher poverty (15.7% vs. 12.7%), lower income ($61,386 vs. $84,046), higher uninsurance (11.8% vs. 9.7%), and older populations (18.7% vs. 14.5% aged 65+). They also live in counties with substantially higher baseline premature mortality (8,935 vs. 6,375 YPLL per 100,000 in 2019), which underscores the health burden of the communities left outside timely access without implying a causal mortality effect. The 30.7 million who improved but did not cross the threshold occupy an intermediate position (poverty 15.8%, income $65,018), confirming that the socioeconomic gradient extends across the entire exclusion spectrum. This gradient is consistent with decades of rural hospital closure and underinvestment that have left the most disadvantaged communities with the thinnest hospital infrastructure onto which AI can be layered. ^13^ Including the instrument-reclassified “lost access” group (14.9 million people who appeared within 30 minutes in 2022 but fell outside in 2024 due to the AHA survey change) would raise the total persistently-outside population from 67.9 to 82.8 million without altering the qualitative equity gradient. The finding identifies specific communities where the next wave of adoption could deliver the largest marginal gains, transforming the distributional finding into an actionable equity agenda. ^14^

### 3.2 Methodological cautions

The SI documents several methodological lessons relevant beyond this study. A short descriptive YPLL context check (SI Section S4) shows that the mortality gradient pre-dates the 2022–2024 AI diffusion window: regressing 2019 YPLL on 2024 AI distance yields a coefficient of +150.0 (*p <* 0.001), close to the 2024 YPLL gradient of +159.2 (*p* = 0.004), reinforcing the interpretation that YPLL is health-burden context rather than evidence of an AI effect. A third survey wave would permit event-study designs that strengthen causal interpretation.

### 3.3 Equity-first diffusion policy

The longitudinal inequality results suggest a practical priority: maximize *marginal coverage gain* and minimize *extreme remoteness*, rather than saturating already-served markets. The socioeconomic profile of the persistently-outside population (Table 4) makes this an equity imperative: communities already facing income deprivation, higher uninsurance, and thinner health-care infrastructure are the same communities that AI diffusion has bypassed. An annual equity-tracking dashboard— maintained by AHRQ or CMS as part of existing quality-reporting infrastructure—could monitor the persistently-outside population, P90 travel distance, and the AI-access Gini to flag whether the access gap is narrowing as diffusion proceeds. Procurement grants and technical assistance can then be targeted to counties where a single new AI-enabled facility would shift the largest number of people below the threshold. ^15^

### 3.4 Limitations and future work

Several limitations are central and deserve explicit discussion.

#### Frame comparability

County-panel and block-group estimates use different analytic universes (Table 5). Headline coverage differs by roughly one percentage point across frames (75.2% county panel vs. 76.1% contiguous-U.S. block groups) because of geographic scope and unit of measurement; inequality trends and transition-group profiles are qualitatively identical across frames.

**Table 5.** Analytic frames and their major results.

| Frame | Units (n) | Key results reported |
| --- | --- | --- |
| County panel (full) | 3,085 counties | Coverage (75.2%), Gini (0.767), P90–P10 gap |
| County cross-section | 3,080 counties | Descriptive YPLL context checks |
| Contiguous-U.S. BG | 236,879 BGs | Maps and contiguous-U.S. coverage/drive-time summaries |
| Full national BG | 240,938 BGs | Table 4 transition profiles, Table S8 Gini bootstrap CIs, operational-only replication |

*Notes:* County panel = counties observed in both waves with valid AI-access measures. Cross-section = panel counties with complete covariates for descriptive YPLL context checks. Contiguous-U.S. BG = block groups in the 48 states + D.C. Full national BG = block groups in the 50 states, D.C., and Puerto Rico.

#### Two-wave panel

The design contains only two periods (2022 and 2024), which limits the analysis to a single pre–post comparison. An event-study specification with three or more waves would allow researchers to verify parallel pre-trends, estimate dynamic effects, and separate adoption timing from calendar-time shocks. Until additional AHA technology waves become available, the 2022–2024 changes should be read as descriptive diffusion patterns rather than causally identified effects.

#### Travel-time proxy

The block-group travel metric uses straight-line distance with a fixed 40 mph speed and 1.3 circuity factor rather than route-level network travel time. Validation against OSRM-routed times for 490 block groups (Section S12) yields mean absolute error (MAE) = 6.7 minutes, 30-minute threshold agreement of 93.5% (sensitivity 97.9%, specificity 83.7%), and a mean bias of −2.0 minutes (proxy understates travel time on average). The dominant error is classifying some truly-outside block groups as within access (false positives for the access class), meaning the proxy under-identifies deserts; the true persistently-outside population is likely larger than reported, making the inequality findings conservative. Accuracy degrades in less-urbanized nonmetro areas (RUCC 7–9: MAE = 13.6 min, misclassification 19.4%) and mountainous terrain (MAE = 13.7 min), with full stratified tables in the SI. The proxy remains the single largest simplification in the spatial design.

#### Access construct and hospital capacity

Our data universe includes all AHA-surveyed hospitals, approximately 30% of which are specialty facilities. A formal exclusion sensitivity (Section S10) confirms that specialty facilities contribute negligibly: excluding them changes coverage by *<*2 pp and the Gini by *<*0.01. As a supply-sensitive benchmark, an E2SFCA index using staffed beds and a 30-minute Gaussian catchment (Section S11) corroborates the nearest-facility findings: the P10 is zero in less-urbanized nonmetro areas in both waves and geographic gradients are concordant across methods. ^10^ This benchmark uses total bed supply because no defensible AI-specific capacity numerator exists at national scale. ^8,9,11^

#### Networked AI and functional access

Geographic proximity to a reporting AI-enabled hospital may not fully capture functional access in networked delivery systems. Some AI capabilities may be centrally hosted by health systems, embedded in shared EHR platforms, or delivered through telehealth and regional referral arrangements. To the extent that non-reporting affiliates benefit from system-level AI infrastructure, nearest-facility measures may overstate access deserts. Conversely, centrally hosted capability may not translate into local clinical workflow integration, staffing capacity, or patient-facing benefit at affiliate hospitals. We therefore interpret our measure as access to locally reported AI deployment, not guaranteed exposure to every AI-enabled service available within a health system.

#### AHA instrument change

The 2022 AHA survey used five binary yes/no AI fields; the 2024 instrument expanded to 14 ordinal implementation items spanning operational and clinical domains. Our crosswalk maps 2022 responses onto a broad “any activity” construct and applies an “active deployment” threshold to 2024 items (analysis stage ≥ 3, corresponding to expanding or fully integrated), but differences in framing, anchoring, and scope could produce measurement-driven adoption changes. In particular, 14.9 million people (409 counties) appeared within 30 minutes of an AI hospital in 2022 but fell outside in 2024; this apparent lost-access group could reflect instrument reclassification rather than service withdrawal (over half lived *<*15 minutes from an AI hospital in 2022). We therefore exclude this group from the “outside, no improvement” category; including it would raise the total outside-both-waves population from 67.9 to 82.8 million while preserving the qualitative equity gradient (Table 4 notes). We mitigate instrument concerns through AI-stage threshold variation (stage ≥ 1 vs. ≥ 3 vs. ≥ 4), an operational-only restriction (63.6% coverage, Gini 0.747), a clinical-only restriction (73.4% coverage, Gini 0.765), and a tight-2022 denominator check requiring at least two affirmative 2022 AI items—all confirming the core findings (see construct-anchor paragraphs above). Hospitals reporting “don’t know” or missing AI-stage responses are coded as non-adopters, a conservative assumption. AHA response rates exceeded 75% in both waves; however, residual nonresponse bias and measurement non-equivalence cannot be ruled out.

#### Observational design

The YPLL analyses are descriptive health-burden context; associations between technology-access distance and premature mortality should not be interpreted as causal effects of AI deployment. ^16,17^ The main contribution of this paper—the longitudinal diffusion, inequality, and health-burden-overlap portrait—does not depend on causal identification of mortality effects.

Future work should extend this design with additional AHA waves, incorporate referral networks and spillovers, and connect diffusion trajectories to hospital-level process outcomes. ^18^ A three-wave design (e.g., 2022, 2024, 2026) would be particularly valuable, enabling event-study specifications and formal pre-trend testing. Natural extensions include propensity-score matching, AI-specific supply-sensitive access metrics, and subgroup-specific inequality measures beyond the Gini.

## 4 Materials and Methods

### 4.1 Design and Data

We integrated two annual waves (2022, 2024) of AHA hospital technology data with county covariates and outcomes (full variable definitions in SI Section S8). County-level analyses include 3,085 counties in the two-period panel (6,170 county-year observations); descriptive YPLL context checks use 3,080 counties with complete covariates. Block-group analyses use two explicitly labeled frames: maps and contiguous-U.S. coverage summaries use 236,879 block groups in the 48 states plus D.C., while transition profiles, Gini bootstrap CIs, and operational-only transition replications use the full national block-group frame of 240,938 block groups in the 50 states, D.C., and Puerto Rico. County YPLL rates are drawn from County Health Rankings, which report multi-year pooled estimates (typically three-year rolling averages ending in the reference year) to stabilize rates in small counties. The temporal lag between YPLL measurement and AHA survey year means the measure is best interpreted as baseline health-burden context rather than an outcome of newly adopted AI.

### 4.2 Access Construction

County exposure and access metrics are based on population-weighted proximity to hospitals with relevant technology flags. Hospital AI flags are constructed from AHA Annual Survey items and harmonized across waves (Table S1). In 2022, we classify a hospital as AI-enabled if it reports using AI/ML for any of five workforce/workflow functions (predicting staffing needs, predicting patient demand, staff scheduling, automating routine tasks, or optimizing administrative/clinical work-flows; fields WFAIPSN, WFAIPPD, WFAISS, WFAIART, WFAIOACW). ^19^ In 2024, we recode the AHA ordinal implementation fields to analysis stages (0=not implementing, 1=exploring, 2=piloting/testing, 3=expanding, 4=fully integrated; “don’t know” and missing values are treated as non-adoption for binary flags) and define adoption under our primary “active deployment” rule as reporting stage ≥ 3 for any of 14 AI items spanning six operational and eight clinical domains (Table S1). ^20^ The crosswalk provides a transparent, policy-interpretable bridge across waves: the 2022 binary yes/no items identify reported AI use in workforce/workflow domains, while the 2024 stage ≥ 3 threshold identifies active deployment under the expanded clinical-plus-operational AI instrument. Because the constructs are not fully equivalent, we interpret changes as changes in measured AI-enabled access under evolving AHA survey instruments, not as a pure longitudinal estimate of the same latent technology construct. We report sensitivity to a broader “any activity” rule (stage ≥ 1, exploring or above) as an upper-bound prevalence scenario. We compute proxy drive time from block-group centroids using haversine distance, 40 mph mean speed, and a 1.3 circuity factor, ^21^ implying an effective 30-minute radius of 15.38 miles. Thirty minutes is a widely used catchment size in health-services spatial accessibility work (e.g., floating-catchment and gravity models) and provides an interpretable emergency-access benchmark. ^8–11^For contiguous-U.S. maps and coverage summaries, block groups are restricted to the 48 states plus D.C.; for full-frame transition profiles, block groups in Alaska, Hawaii, and Puerto Rico are retained. Distances are computed to the nearest qualifying hospital in the full geocoded AHA hospital frame (50 states plus D.C.), regardless of state boundaries. All-hospital distances and inequality are nearly unchanged between waves (Table 1), confirming that the AI-specific results are not driven by general hospital churn.

The distance-to-minutes conversion is a constant rescaling, so continuous-distance inequality metrics (Lorenz curve, Gini) are invariant to speed and circuity assumptions. Threshold-coverage rates shift with parameterization: under plausible ranges (30– 50 mph; circuity 1.2–1.4), 2024 coverage spans approximately 64–85%. We therefore report the outside-both-waves population for the prespecified 40 mph, 1.3 circuity, 30-minute benchmark: 82.8 million people before excluding the 14.9 million instrument-reclassified lost-access group, and 67.9 million after that exclusion. Under all parameterizations, coverage increased between waves while the Gini rose.

This proxy is the single largest simplification in the spatial design. We validated it by comparing circuity-adjusted Euclidean estimates against OSRM network-routed times for a stratified random sample of 500 block groups (490 successfully routed). Overall concordance is strong (MAE = 6.7 min, RMSE = 13.1 min; 30-minute threshold agreement = 93.5%), with a mean bias of −2.0 minutes (the proxy understates travel time on average, meaning it overstates access; the dominant misclassification is placing some truly-outside block groups within the 30-minute threshold, so the true desert population is likely larger than reported). Accuracy degrades in less-urbanized nonmetro areas (RUCC 7–9) and mountainous areas (MAE = 13.6 and 13.7 min, respectively), consistent with prior transportation research on circuity variation. ^21^ Full stratified error metrics by RUCC, census division, and terrain appear in SI Section S12.

### 4.3 Health-Burden Context

The main text profiles baseline 2019 county YPLL across access-transition groups. SI Section S4 reports descriptive county-level checks, including the pre-diffusion YPLL comparison used to assess whether the mortality gradient predates the 2022–2024 AI expansion window. These checks characterize baseline health burden in communities left outside timely AI access; they are not interpreted as causal outcome effects of AI deployment.

### 4.4 Comparability Notes

County-panel and block-group estimates use different analytic frames (see Limitations, Table 5). All within-frame longitudinal comparisons use fixed universes; table and figure captions identify the applicable frame.

## Data Availability

Hospital-level data from the AHA Annual Survey are available under license from the American Hospital Association (https://www.ahadata.com). Aggregated county-level analytic datasets, all analysis code, and pipeline reproducibility scripts are publicly available at https://github.com/undergroundbreakfast/Scientific_Paper (tag v1.0.0); a permanent archival copy will be deposited with Zenodo upon journal acceptance (DOI to be assigned). Block-group-level Census population data are from the U.S. Census Bureau (2020 Decennial Census). County Health Rankings data are publicly available at https://www.countyhealthrankings.org.

## Ethics Statement

This study used deidentified institutional records and aggregate geographic data and did not involve human-subjects interaction; IRB review was not required.

## Author Contributions

A.J. designed research, performed research, analyzed data, and wrote the paper; D.G. and T.H. designed research and edited the paper.

## Competing Interests

The authors declare no competing interests.

## Supplementary Information

Supplementary Materials for: Rapid Diffusion, Persistent Deserts

### S1 AI Taxonomy and Wave Crosswalk

To clarify how hospital AI is measured and harmonized across waves, Table S1 lists the AHA survey items and response scales used to construct our AI flags.

**Table S1.** Hospital AI survey items used to construct AI flags in 2022 and 2024.

| Wave | Field | AHA item description | Response |
| --- | --- | --- | --- |
| **Hospital AI — Operational domains (2022 & 2024)** |  |  |  |
| 2022 | WFAIPSN | AI or machine learning — predicting staffing needs | 1=yes, 0=no |
| 2022 | WFAIPPD | AI or machine learning — predicting patient demand | 1=yes, 0=no |
| 2022 | WFAISS | AI or machine learning — staff scheduling | 1=yes, 0=no |
| 2022 | WFAIART | AI or machine learning — automating routine tasks | 1=yes, 0=no |
| 2022 | WFAIOACW | AI or machine learning — optimizing administrative and clinical workflows | 1=yes, 0=no |
| 2024 | AIOPEF | Optimizing operational efficiency — operation AI implementation | 0–4 recoded |
| 2024 | AIPATD | Patient flow and demand forecasting — operation AI implementation | 0–4 recoded |
| 2024 | AIREVC | Revenue cycle management — operation AI implementation | 0–4 recoded |
| 2024 | AISCOP | Supply chain optimization — operation AI implementation | 0–4 recoded |
| 2024 | AISSWM | Staff scheduling and workforce management — operation AI implementation | 0–4 recoded |
| 2024 | AITIOT | Other — operation AI implementation | 0–4 recoded |
| **Clinical AI (2024 only)** |  |  |  |
| 2024 | CLINAI | Clinical decision support tools | 0–4 recoded |
| 2024 | CLNOAI | Other clinical AI implementation | 0–4 recoded |
| 2024 | DIAGAI | AI-assisted diagnostics | 0–4 recoded |
| 2024 | PAPCAI | Predictive analytics for patient care | 0–4 recoded |
| 2024 | PCOEAI | Patient communication and education | 0–4 recoded |
| 2024 | PDMRAI | Resource allocation during emergencies | 0–4 recoded |
| 2024 | POPHAI | Population health management | 0–4 recoded |
| 2024 | SURGAI | AI-assisted surgery | 0–4 recoded |

Notes: For 2024 AI items, text responses are recoded to analysis stages: 0=not implementing, 1=exploring, 2=piloting/testing, 3=expanding, and 4=fully integrated; “do not know” and missing values are treated as non-adoption for binary flags. In the main analysis we define 2024 “active deployment” as stage $\geq 3$ (expanding or fully integrated) on any of the 14 operational and clinical AI items, and “any activity” as stage $\geq 1$ (exploring or above; Materials and Methods). The 2024 instrument expanded the AI construct to include both operational and clinical domains; we include all 14 fields to capture the full scope of hospital AI implementation. Item wording and codes come from AHA/Health Forum annual survey file-layout documents for FY2022 and FY2024.<sup>19,20</sup>

### S2 Census-Division Heterogeneity

**Table S2.** Census-division changes in mapped hospital AI adoption, 2022–2024.

| Div. | Division | Region | States Included | 2022 Mapped Mean | 2024 Mapped Mean | Diff |
| --- | --- | --- | --- | --- | --- | --- |
| 7 | West South Central | South | Arkansas, Louisiana, Oklahoma, Texas | 0.0625 | 0.1153 | +0.0528 |
| 4 | West North Central | Midwest | Iowa, Kansas, Minnesota, Missouri, Nebraska, North Dakota, South Dakota | 0.1119 | 0.1534 | +0.0415 |
| 8 | Mountain | West | Arizona, Colorado, Idaho, Montana, Nevada, New Mexico, Utah, Wyoming | 0.0749 | 0.1059 | +0.0311 |
| 9 | Pacific | West | Alaska, California, Hawaii, Oregon, Washington | 0.0573 | 0.0854 | +0.0281 |
| 6 | East South Central | South | Alabama, Kentucky, Mississippi, Tennessee | 0.0793 | 0.1003 | +0.0210 |
| 5 | South Atlantic | South | DE, DC, FL, GA, MD, NC, SC, VA, WV | 0.0340 | 0.0496 | +0.0156 |
| 3 | East North Central | Midwest | Illinois, Indiana, Michigan, Ohio, Wisconsin | 0.0965 | 0.1042 | +0.0077 |
| 1 | New England | Northeast | CT, ME, MA, NH, RI, VT | 0.1407 | 0.1472 | +0.0065 |
| 2 | Middle Atlantic | Northeast | New Jersey, New York, Pennsylvania | 0.1765 | 0.1756 | -0.0010 |

Notes: Values are division-level means of mapped hospital AI adoption share (0–1). Because 2024 AHA AI items are more extensive than 2022, estimates use the manuscript crosswalk as a transparent bridge across evolving survey instruments rather than as a fully equivalent latent construct. Divisions ordered by 2022–2024 change.

### S3 AI-Flag Sensitivity

We tested sensitivity of the 2024 results to alternative definitions of the AI-enabled hospital flag. The main specification uses the “active deployment” threshold (analysis stage ≥ 3, expanding or fully integrated, on any of the 14 AI implementation items), while the looser “any activity” threshold (stage ≥ 1, exploring or above) serves as an upper-bound scenario. In addition, we tested a construct-validity restriction that limits the 2024 flag to the six operational items only (AIOPEF, AIPATD, AIREVC, AISCOP, AISSWM, AITIOT), excluding the eight clinical AI columns that have no direct 2022 counterpart.

**Fig. S1.**
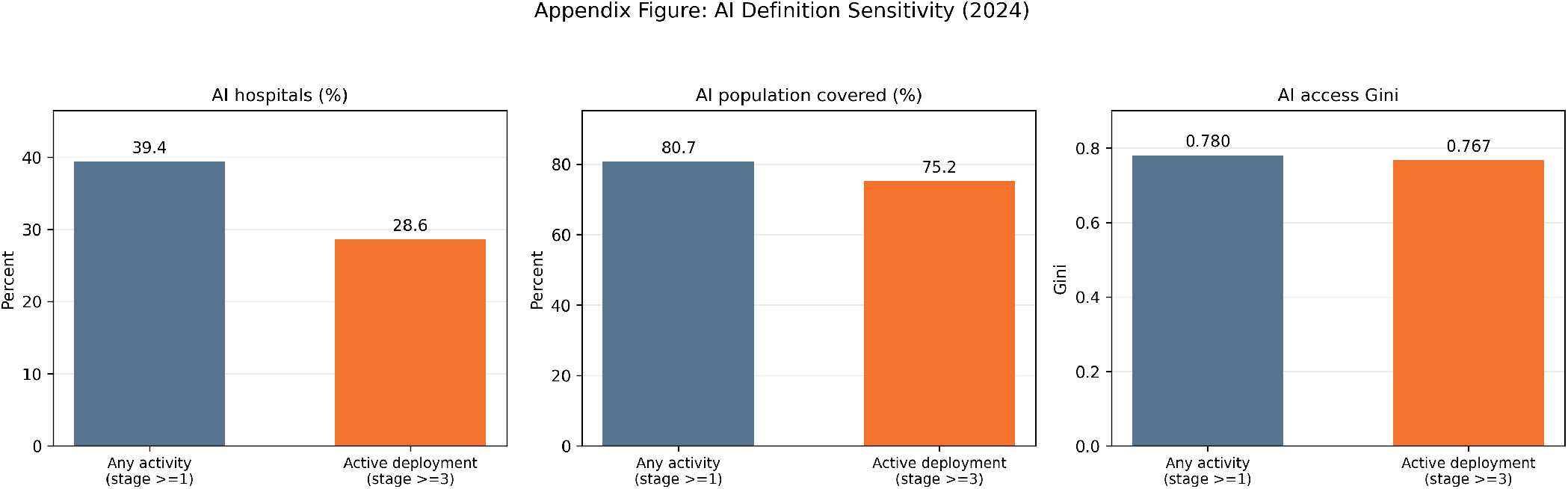
2024 AI-flag sensitivity. *Any activity* (stage ≥ 1, exploring or above) yields 39.4% prevalence and 80.7% coverage (Gini 0.780); the stricter *active deployment* (stage ≥ 3, expanding or fully integrated) threshold yields 28.6% prevalence and 75.2% coverage (Gini 0.767). Persistent inequality in either specification supports reporting *active deployment* as the conservative primary definition.

Table S3 reports the operational-only restriction. Limiting the AI flag to six operational items at stage ≥ 3 reduces the qualifying hospital count from 1,743 to 1,091 (18.0% of all hospitals), with coverage falling from 75.2% to 63.6% in the full frame. The 11.6 percentage-point gap shows that clinical AI adopters contribute meaningfully to measured spatial access. Crucially, the Gini coefficient remains high under the operational-only definition (0.747 full frame vs. 0.767 for all 14 items), confirming that the core inequality finding is robust to construct scope.

**Table S3.** Operational-only construct-validity sensitivity (2024). Comparison of the full 14-item hospital AI definition versus a restriction to the 6 operational items only, both at the active-deployment threshold (stage ≥ 3, expanding or fully integrated).

| Metric | Hospital AI (14 items) | Operational only (6 items) | Difference |
| --- | --- | --- | --- |
| AI hospitals (*n*) | 1,743 | 1,091 | −652 |
| Hospital prevalence (%) | 28.6 | 18.0 | −10.6 |
| Coverage, full frame (%) | 75.2 | 63.6 | −11.6 |
| Coverage, contiguous U.S. (%) | 76.1 | 64.4 | −11.7 |
| Gini, full frame | 0.767 | 0.747 | −0.020 |

*Notes:* The six operational items are AIOPEF, AIPATD, AIREVC, AISCOP, AISSWM, and AITIOT. The eight excluded clinical items are CLINAI, CLNOAI, DIAGAI, PAPCAI, PCOEAI, PDMRAI, POPHAI, and SURGAI. Of the 1,743 hospitals meeting the 14-item active-deployment threshold, 1,737 have valid coordinates and enter the spatial analysis; of those, 911 have both operational and clinical AI at stage $\geq 3$ , 180 have operational only, and 646 have clinical AI only.

Table S4 reports a stricter deployment threshold sensitivity. Raising the bar from stage ≥ 3 (expanding or fully integrated) to stage ≥ 4 (fully integrated only) halves the AI-enabled hospital count from 1,743 to 859 (14.1%), and 30-minute population coverage drops from 75.2% to 58.5%—a 16.7 percentage-point reduction. The Gini coefficient falls slightly to 0.732 (from 0.767), indicating that the most advanced deployers are somewhat more evenly distributed than the broader set, though inequality remains high. The coverage drop is substantially larger than under the operational-only restriction (− 11.6 pp) or the clinical-only restriction (− 0.4 pp), suggesting that many hospitals qualifying at stage ≥ 3 are expanding but not yet fully integrated. Taken together, the threshold and construct variants span a wide range of prevalence (14.1–39.4%) and coverage (58.5–80.7%), yet all produce Gini coefficients above 0.73, confirming that the main inequality finding is robust to how “AI-enabled” is defined.

**Table S4.**
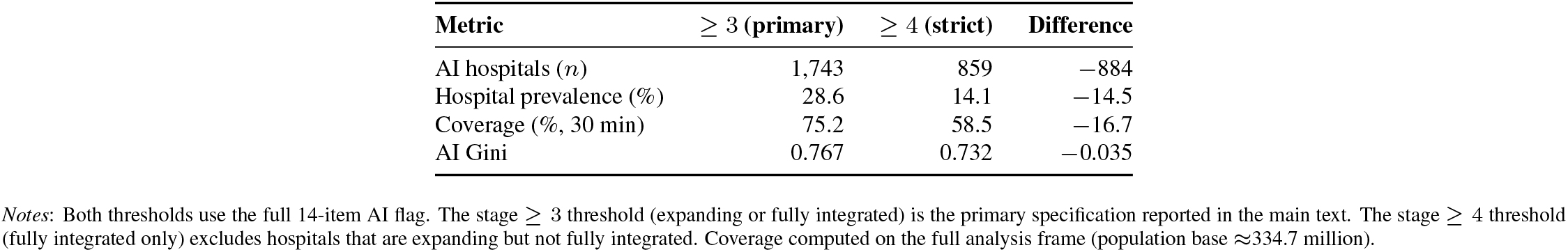
Deployment-threshold sensitivity (2024). Comparison of the primary stage ≥ 3 threshold (expanding or fully integrated) with a stricter stage ≥ 4 threshold (fully integrated only).

**Table S5.** Access-threshold sensitivity for transition profiles. Population counts are full-frame millions. “Outside both” excludes the apparent lost-access group.

| Threshold | Coverage 2022 | Coverage 2024 | Newly within | Outside both | Improved, still outside | Outside, no improvement | Apparent lost access |
| --- | --- | --- | --- | --- | --- | --- | --- |
| 30 min | 221.7 | 251.9 | 45.1 | 67.9 | 30.7 | 37.2 | 14.9 |
| 45 min | 250.8 | 279.2 | 42.8 | 41.1 | 18.2 | 22.8 | 14.5 |
| 60 min | 270.3 | 295.4 | 37.2 | 27.2 | 11.6 | 15.6 | 12.1 |

*Notes:* Counts are population totals in millions, computed from the full block-group transition frame (334.7 million residents). “Outside both” equals block groups outside the threshold in both 2022 and 2024 and excludes block groups that appeared inside the threshold in 2022 but outside in 2024. Longer thresholds reduce the absolute outside-both-waves population, but a substantial outside-both group remains under 45- and 60-minute catchments.

**Table S6.** Tight 2022 AI-use sensitivity. The tight 2022 specification requires at least two affirmative workforce/workflow AI items in 2022; the 2024 comparison uses the primary stage ≥ 3 active-deployment flag.

| Specification | AI hospitals | 30-min coverage | Gini | Outside both vs. 2024 |
| --- | --- | --- | --- | --- |
| 2022 main: $\geq 1$ affirmative workforce/workflow item | 1,118 | 221.7M (66.2%) | 0.739 | 67.9M |
| 2022 tight: $\geq 2$ affirmative workforce/workflow items | 878 | 207.2M (61.9%) | 0.743 | 70.5M |
| 2024 primary: stage $\geq 3$ on any active AI item | 1,737 | 251.9M (75.3%) | 0.767 | — |

*Notes:* Hospital counts in this table are the geocoded spatial-analysis counts used for coverage and Gini calculations; they are lower than the full survey counts in Table 1 by one hospital in 2022 and six hospitals in 2024 because spatial coverage requires usable coordinates. Coverage and Gini use the full block-group analysis frame. Under the tight 2022 denominator, the 2022-to-2024 transition profile includes 57.1 million newly within 30 minutes, 70.5 million outside both waves, and 12.4 million in the apparent lost-access group. Of the 70.5 million outside both waves, 40.5 million improved but remained outside and 30.0 million had no travel-time improvement.

### S4 Health-Burden Context: Descriptive YPLL Checks

This section keeps YPLL in a narrow role: descriptive health-burden context for the access gradients reported in the main text. The estimates below do not identify an effect of AI deployment on mortality.

#### Log-distance context models

County-level models relate log distance to the nearest AI-enabled hospital to Years of Potential Life Lost before age 75 (YPLL), controlling for social/economic factors, health behaviors, physical environment, log population, and census-division fixed effects. The 2024 coefficient is positive and statistically significant; the 2022 coefficient is positive but not significant (Table S7). These coefficients should be read as descriptive mortality gradients associated with geography, not as causal effects of AI deployment.

#### Pre-AI falsification

Replacing the outcome with 2019 YPLL (County Health Rankings, pooled ≈2016–2018) while retaining 2024 AI distance yields a positive coefficient (+150.0, *p <* 0.001; Table S7). This confirms that the spatial mortality gradient pre-dates the 2022–2024 diffusion window: counties far from 2024 AI hospitals already had higher premature mortality before those hospitals adopted AI. The appropriate interpretation is therefore burden overlap, not AI impact.

**Table S7.** Descriptive YPLL context checks. Positive coefficients indicate that greater distance to AI-enabled hospitals is associated with higher county YPLL.

| Model | AI-distance coefficient (p-value) | R<sup>2</sup> |
| --- | --- | --- |
| 2022 YPLL on 2022 log-distance | +40.1 (0.353) | 0.677 |
| 2024 YPLL on 2024 log-distance | +159.2 (0.004) | 0.710 |
| 2019 YPLL on 2024 log-distance (pre-AI falsification) | +150.0 (< 0.001) | 0.674 |

Notes: The 2022 and 2024 models use complete-covariate county samples and include census-division fixed effects, social/economic factors, health behaviors, physical environment, and log population. The 2019 falsification sample has $n = 2,899$ because 181 counties lack 2019 YPLL data. Results are descriptive and should not be interpreted as causal mortality evidence.

### S5 Bootstrap Confidence Intervals for Inequality Metrics

To assess the statistical significance of inequality changes, we computed county-clustered bootstrap confidence intervals for the population-weighted Gini coefficient of block-group distances. In each of *B* = 500 iterations, counties were resampled with replacement (retaining all block groups within each sampled county), and the Gini was recomputed on the resampled population. Paired resampling—drawing the same set of counties for both waves in each iteration—ensures that the change-in-Gini CI reflects within-county variation rather than cross-county sampling noise. Results are reported for AI-enabled hospitals and all hospitals as the baseline geography (Table S8).

The AI Gini increase of 0.028 (from 0.739 to 0.767) is statistically significant at the 5% level (95% CI [0.016, 0.038]), confirming that the main inequality finding is not attributable to compositional instability. The all-hospital Gini is unchanged between waves (change ≈ 0; CI straddles zero), indicating that the inequality increase is specific to AI deployment geography.

**Table S8.** Bootstrap confidence intervals for population-weighted Gini coefficients of block-group distances to nearest technology-enabled hospital.

| Technology | Year | Gini | 95% CI | Bootstrap Mean |
| --- | --- | --- | --- | --- |
| All hospitals | 2022 | 0.832 | [0.793, 0.862] | 0.830 |
| All hospitals | 2024 | 0.831 | [0.791, 0.861] | 0.830 |
| AI hospitals | 2022 | 0.739 | [0.712, 0.764] | 0.737 |
| AI hospitals | 2024 | 0.767 | [0.736, 0.795] | 0.766 |
| Change in Gini (2022→2024) |  |  |  |  |
| All hospitals | — | −0.0001 | [−0.0007, +0.0005] | — |
| AI hospitals | — | +0.028 | [+0.016, +0.038] | — |

Notes: County-clustered bootstrap with $B = 500$ iterations. Resampling unit = county; all block groups within each sampled county are retained. Paired resampling ensures that the same set of counties is drawn for both waves in each iteration. Universe: 240,938 block groups across all sampled counties. The AI Gini increase is the only change that is statistically significant at the 5% level.

### S6 Alternative Inequality Metrics

Table S9 reports Atkinson and Theil indices as distributional robustness checks alongside the population-weighted Gini coefficient. The AI access-distance distribution becomes more unequal under all three metrics between 2022 and 2024, while the all-hospital baseline remains essentially unchanged.

**Table S9.** Alternative inequality metrics for population-weighted access-distance distributions.

| Access type | Gini | | Atkinson ($\epsilon = 0.5$) | | Theil T | |
| --- | --- | --- | --- | --- | --- | --- |
|  | 2022 | 2024 | 2022 | 2024 | 2022 | 2024 |
| AI hospitals | 0.739 | 0.767 | 0.477 | 0.526 | 1.432 | 1.730 |
| All hospitals | 0.832 | 0.831 | 0.667 | 0.667 | 2.717 | 2.716 |

*Notes:* Metrics are computed on block-group distances to the nearest qualifying hospital and weighted by 2020 block-group population. Higher values indicate greater inequality in access-distance burden. Exact co-location distances are represented by the pipeline’s 0.1-mile floor before distributional calculations.

### S7 Operational-Only Transition-Group Replication

To isolate the contribution of construct expansion (the eight clinical AI items added in 2024) from true operational AI diffusion, we replicated the longitudinal transition-group analysis (Table 4 in the main text) under the operational-only restriction, limiting the 2024 AI flag to the six operational items at the active-deployment threshold (stage ≥ 3, expanding or fully integrated). Table S10 compares the main (14-item) and operational-only (6-item) transition groups.

#### Note on group definitions

The “outside, no improvement + lost access” population in this table (52.2 million under the main definition) includes the full pre-adjustment universe—i.e., the 37.2 million in the “outside, no improvement” group *plus* the 14.9 million in the “lost-access” artifact group attributed to instrument change (see AHA Instrument Change in main-text Limitations). Table 4 in the main text separates the “improved, still outside” group (30.7 million) from the “outside, no improvement” group (37.2 million) after excluding the lost-access artifact. The operational-only comparison uses the unadjusted universe in both columns to ensure a like-for-like contrast across AI definitions.

**Table S10.** Transition-group comparison: main (14-item) vs. operational-only (6-item) AI definitions.

| Transition Group | Main (million) | Op-Only (million) | Diff (million) | % Change |
| --- | --- | --- | --- | --- |
| Persistently within 30 min | 206.8 | 185.1 | −21.7 | −10.5% |
| Newly within 30 min | 45.1 | 27.9 | −17.2 | −38.2% |
| Improved, still outside | 30.7 | 28.8 | −1.9 | −6.2% |
| Outside, no improvement + lost access | 52.2 | 93.0 | +40.8 | +78.3% |

*Notes:* Main definition uses all 14 AI items (6 operational + 8 clinical) at the active-deployment threshold (stage $\geq 3$ , expanding or fully integrated) for the 2024 wave. Operational-only restricts to 6 operational items (AIOPEF, AIPATD, AIREVC, AISCOP, AISSWM, AITIOT) at the same stage $\geq 3$ threshold. The 2022 AI flag is identical across both definitions (5 binary operational items). Population totals are based on the full block-group universe (334.7 million). The outside/no-improvement plus lost-access group nearly doubles under the operational-only restriction relative to the same unadjusted denominator, confirming that clinical AI items substantially expand measured 2024 coverage.

The operational-only restriction sharply reduces the “newly within” population (−38.2%) and correspondingly increases the outside/no-improvement plus lost-access group (+78.3% relative to the same unadjusted denominator), demonstrating that a substantial share of the measured 2022–2024 coverage expansion is attributable to the eight clinical AI items introduced in the 2024 instrument. Against the main adjusted persistently-outside benchmark of 67.9 million, the operational-only estimate of 93.0 million represents a 37% increase. The “improved but still outside” group is relatively stable (−6.2%), indicating that distance improvements among communities outside 30 minutes are largely driven by operational AI diffusion rather than clinical AI additions. These results provide the strongest available evidence that the core inequality finding is not an artifact of instrument expansion: even under the most conservative common-item restriction—where 2024 operational-only prevalence (18.0%) is virtually unchanged from the 2022 headline (18.3%)—spatial inequality persisted (Gini 0.747). The robust finding is persistent inequality across construct definitions, not necessarily rapid common-item operational AI diffusion.

### S8 Variable Definitions

Table S11 defines the covariates used in descriptive YPLL context checks and access analyses.

### S9 Clinical-Only 2024 AI Sensitivity

The preceding operational-only analysis (Section S7) guards against construct expansion by restricting to items present in both survey waves. A complementary concern is whether the access results are driven almost entirely by the *clinical* AI domains added in 2024 rather than by the full 14-item construct. To address this, we re-estimate access and inequality metrics restricting the 2024 AI flag to the eight *clinical* items only: clinical decision support (CLINAI), other clinical AI (CLNOAI), AI-assisted diagnostics (DIAGAI), predictive analytics for patient care (PAPCAI), patient communication and education (PCOEAI), resource allocation during emergencies (PDMRAI), population health management (POPHAI), and AI-assisted surgery (SURGAI)—each at the active-deployment threshold (stage ≥ 3, expanding or fully integrated).

**Table S11.**
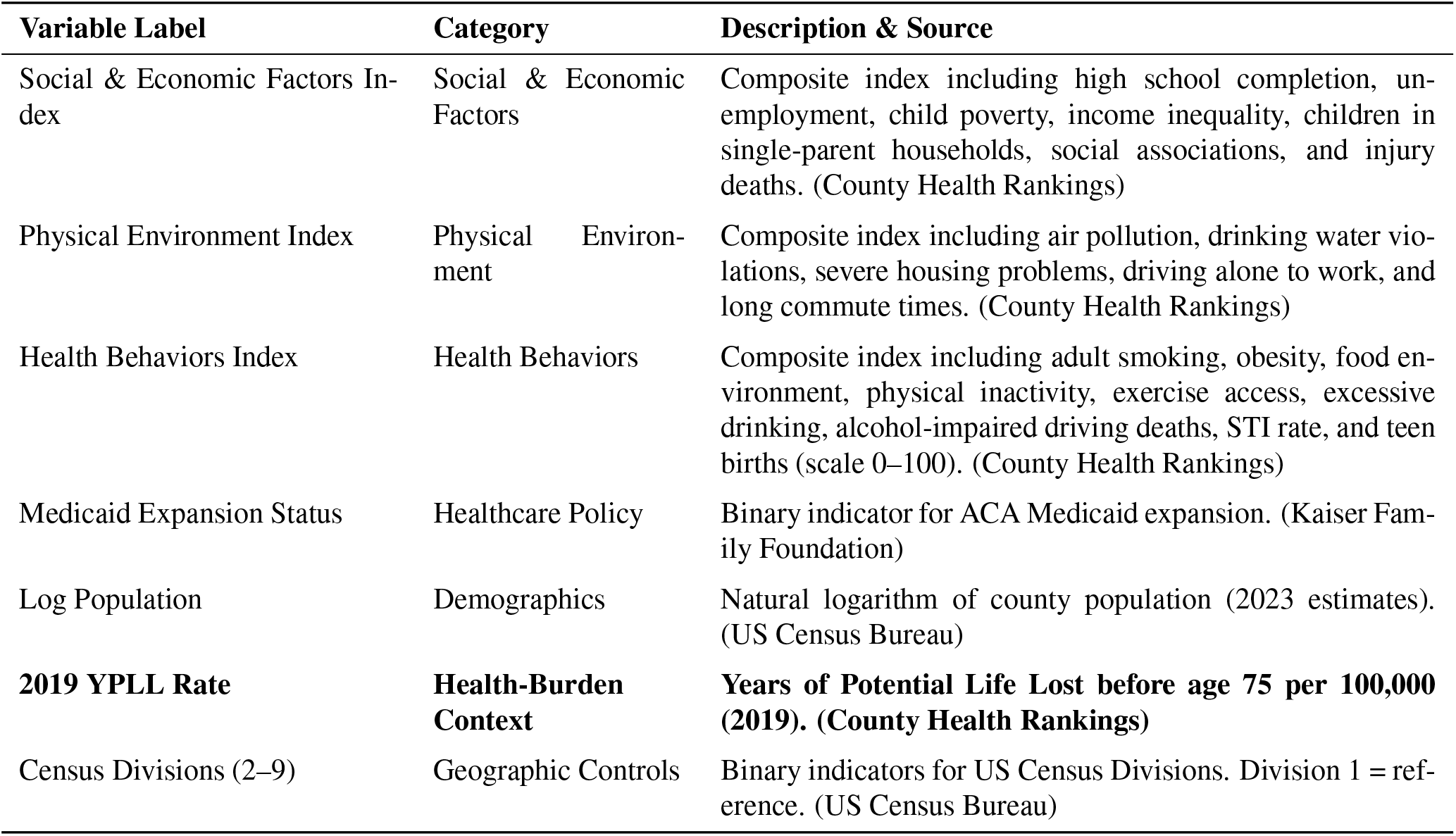
Covariate Definitions and Data Sources.

Table S12 compares the clinical-only and full-definition results across access metrics. The clinical-only flag identifies 1,557 hospitals (25.6%), vs. 1,743 (28.6%) under the full definition. Population coverage is slightly lower (73.4% vs. 75.2%), and the Gini coefficient is nearly unchanged (0.765 vs. 0.767), confirming that the inequality finding is not an artifact of including operational AI items in the broader 14-item construct.

**Table S12.** Clinical-only vs. full-definition AI sensitivity: 2024 cross-section.

| Metric | Clinical-Only (8 items) | Full Definition (14 items) |
| --- | --- | --- |
| AI-enabled hospitals | 1,557 (25.6%) | 1,743 (28.6%) |
| Population coverage (30 min) | 73.4% | 75.2% |
| Population-weighted Gini | 0.765 | 0.767 |

Notes: Clinical-only flag requires stage $\geq 3$ (expanding or fully integrated) on any of the eight clinical AI items (CLINAI, CLNOAI, DIAGAI, PAPCAI, PCOEAI, PDMRAI, POPHAI, SURGAI). Full definition requires stage $\geq 3$ on any of 14 items (6 operational + 8 clinical). Both use the active-deployment threshold. Hospital counts are survey totals; spatial coverage uses the geocoded subset (1,737 of 1,743 full-definition hospitals have valid coordinates).

### S10 General Acute-Care Hospital Exclusion Sensitivity

The main analysis includes all AHA-surveyed hospitals reporting technology data, regardless of facility type. To test whether specialty facilities (e.g., psychiatric, long-term care, rehabilitation) influence the core results, we re-estimate all access and inequality metrics after restricting the hospital universe to general acute-care facilities only, classified via AHA service-type fields. Table S13 reports the comparison.

Excluding specialty hospitals removes 1,816 facilities in 2022 (29.8% of all hospitals) and 1,871 in 2024 (30.8%), reducing the AI-enabled hospital count from 1,119 to 985 (−12.0%) and from 1,743 to 1,567 (−10.1%). Despite this substantial reduction in the hospital universe, access metrics are minimally affected: 30-minute AI coverage declines by only 1.7 percentage points in 2022 and 0.4 pp in 2024, while the population-weighted Gini changes by less than 0.01 in both waves (0.739 to 0.731; 0.767 to 0.763). The P90–P10 distance gap and mean population-weighted distance are similarly stable. These results confirm that specialty facilities contribute negligibly to AI spatial access patterns, because the nearest AI-enabled hospital for almost all block groups is a general acute-care facility.

**Table S13.** Specialty-hospital exclusion sensitivity: all hospitals vs. general acute care only.

| Metric | 2022 |  |  | 2024 |  |  |
| --- | --- | --- | --- | --- | --- | --- |
|  | All | Gen. acute | Diff. | All | Gen. acute | Diff. |
| Total hospitals | 6,095 | 4,279 | −1,816 | 6,076 | 4,205 | −1,871 |
| AI hospitals (*n*) | 1,119 | 985 | −134 | 1,743 | 1,567 | −176 |
| AI prevalence (%) | 18.3 | 23.0 | +4.7 | 28.6 | 37.3 | +8.7 |
| AI coverage (%; 30 min) | 66.2 | 64.5 | −1.7 | 75.2 | 74.8 | −0.4 |
| AI Gini | 0.739 | 0.731 | −0.008 | 0.767 | 0.763 | −0.004 |
| P90–P10 gap (miles) | 49.8 | 52.6 | +2.8 | 33.2 | 33.4 | +0.2 |
| Mean distance (miles) | 28.9 | 29.7 | +0.8 | 23.2 | 23.4 | +0.2 |

*Notes:* General acute-care hospitals classified via AHA service-type field (“General medical and surgical”); all other facility types (psychiatric, rehabilitation, long-term care, children’s, etc.) excluded. Coverage and Gini computed on the full analysis frame (population base $\approx 334.7$ million). AI prevalence rises under general acute care only because the denominator shrinks (specialty hospitals are less likely to report AI deployment). Hospital counts are survey totals; spatial metrics use the geocoded subset.

### S11 Enhanced Two-Step Floating Catchment Area (E2SFCA) Benchmark

The main analysis uses nearest-facility distance and threshold coverage, which are transparent and interpretable but do not account for hospital capacity or competition. As a supply-sensitive benchmark, we computed an Enhanced Two-Step Floating Catchment Area (E2SFCA) accessibility index for all block groups in both waves, using staffed hospital beds as the capacity numerator and a 30-minute Gaussian distance-decay catchment. ^10^ The E2SFCA score represents the effective bed supply accessible to each block group, accounting for competition from surrounding populations.

Table S14 reports population-weighted E2SFCA access by RUCC group. The key patterns corroborate the nearest-facility findings: (i) median access declines from metro to less-urbanized nonmetro areas in both waves; (ii) the P10 is zero in less-urbanized nonmetro areas in both waves, meaning some block groups have no hospital beds within their 30-minute catchment—consistent with the access-desert finding; and (iii) access is essentially stable between waves overall (weighted mean 0.00274 in 2022 vs. 0.00271 in 2024), with slight declines in both nonmetro bins. The high P90 in the less-urbanized nonmetro bin reflects small rural hospitals serving small catchment populations, producing high beds-per-person ratios for the few block groups near them. This confirms that the rural access problem is one of extensive margin (zero access) rather than intensive margin (inadequate capacity where access exists).

Table S15 reports E2SFCA access by census division. The East South Central division has the highest mean access (driven by high bed density in smaller metros), while the Pacific has the lowest. The pattern is broadly consistent with the nearest-facility distance rankings reported in the main text and underscores that supply-sensitive and nearest-facility approaches yield concordant geographic gradients for this application.

**Table S14.** E2SFCA access index by RUCC group (bed-weighted, 30-min Gaussian catchment).

| RUCC group | Year | Pop-wtd mean | P10 | P50 | P90 |
| --- | --- | --- | --- | --- | --- |
| Metro (1–3) | 2022 | 0.00278 | 0.00082 | 0.00257 | 0.00493 |
| Metro (1–3) | 2024 | 0.00277 | 0.00080 | 0.00256 | 0.00494 |
| Nonmetro, more urban (4–6) | 2022 | 0.00238 | 0.00019 | 0.00163 | 0.00519 |
| Nonmetro, more urban (4–6) | 2024 | 0.00229 | 0.00017 | 0.00155 | 0.00491 |
| Nonmetro, less urban (7–9) | 2022 | 0.00263 | 0.000 | 0.00157 | 0.00608 |
| Nonmetro, less urban (7–9) | 2024 | 0.00254 | 0.000 | 0.00150 | 0.00600 |

*Notes:* E2SFCA index computed using staffed beds (BSC) as the capacity numerator and a Gaussian distance-decay function within a 30-minute catchment (haversine distance, 40 mph, 1.3 circuitry factor). Higher values indicate greater effective bed supply per capita. P10 = 0 for the less-urbanized nonmetro bin indicates that at least 10% of that population has zero hospital beds within their 30-minute catchment. RUCC 4–6 and 7–9 are rurality-size bins rather than ERS metro-adjacency bins. *n* = 240,938 block groups; population base $\approx 334.7$ million. Hospital universe includes all AHA-surveyed hospitals regardless of technology flag.

### S12 Proxy Travel-Time Validation

To quantify the accuracy of the circuity-adjusted Euclidean proxy used throughout this study, we compared proxy travel times against OpenStreetMap (OSM) network-routed times via the OSRM engine for a stratified random sample of 500 block groups (490 successfully routed). Table S16 reports overall error metrics; Tables S17–S18 stratify by USDA Rural-Urban Continuum Code (RUCC) and census division; Table S19 reports 30-minute threshold classification accuracy by terrain type.

**Table S15.** E2SFCA access index by census division (bed-weighted, 30-min Gaussian catchment).

| Division | 2022 |  |  | 2024 |  |  |
| --- | --- | --- | --- | --- | --- | --- |
|  | Mean | P10 | P90 | Mean | P10 | P90 |
| East South Central | 0.00354 | 0.00056 | 0.00738 | 0.00352 | 0.00053 | 0.00750 |
| West North Central | 0.00329 | 0.00046 | 0.00644 | 0.00322 | 0.00045 | 0.00627 |
| West South Central | 0.00296 | 0.00054 | 0.00547 | 0.00297 | 0.00054 | 0.00556 |
| Middle Atlantic | 0.00295 | 0.00124 | 0.00462 | 0.00292 | 0.00119 | 0.00456 |
| East North Central | 0.00281 | 0.00077 | 0.00489 | 0.00274 | 0.00072 | 0.00488 |
| South Atlantic | 0.00273 | 0.00070 | 0.00498 | 0.00274 | 0.00068 | 0.00501 |
| New England | 0.00271 | 0.00096 | 0.00486 | 0.00269 | 0.00093 | 0.00481 |
| Mountain | 0.00240 | 0.00038 | 0.00403 | 0.00244 | 0.00038 | 0.00408 |
| Pacific | 0.00215 | 0.00074 | 0.00345 | 0.00209 | 0.00068 | 0.00344 |

Notes: Population-weighted mean E2SFCA access. Divisions ordered by 2022 mean access (descending). Same computation as Table S14. “Unknown” division ( $n = 2,505$ block groups, 3.3M population) omitted; these block groups have zero E2SFCA access in both waves.

**Table S16.**
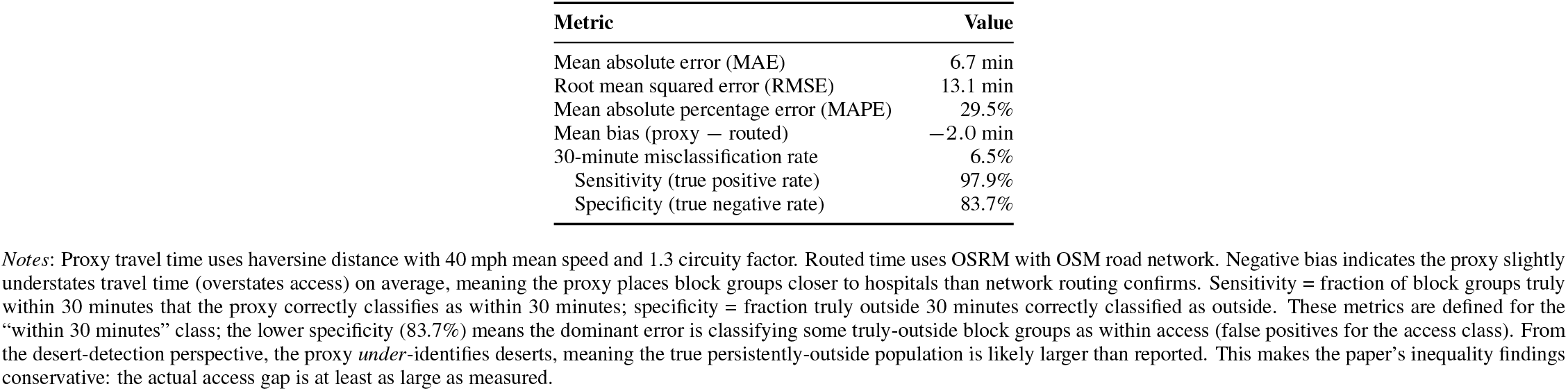
Proxy validation: overall error metrics (*n* = 490).

**Table S17.** Proxy validation stratified by RUCC group.

| RUCC group | $n$ | MAE (min) | RMSE (min) | Bias (min) | Misclass. rate | Sensitivity |
| --- | --- | --- | --- | --- | --- | --- |
| Metro (1–3) | 409 | 6.0 | 12.0 | –1.5 | 5.9% | 98.4% |
| Nonmetro, more urban (4–6) | 45 | 8.7 | 18.0 | –2.2 | 2.2% | 93.8% |
| Nonmetro, less urban (7–9) | 31 | 13.6 | 19.3 | –8.3 | 19.4% | 75.0% |
| Missing/unknown RUCC | 5 | — | — | — | — | — |

Notes: RUCC codes from USDA Economic Research Service (2023). Metro = counties in metro areas of any size (RUCC 1–3); RUCC 4–6 and 7–9 are rurality-size bins rather than metro-adjacency bins. ERS adjacency cuts across codes 4/6/8 versus 5/7/9. Five successfully routed block groups had missing/unknown RUCC and are listed separately so strata sum to the overall validation sample ( $n = 490$ ). The proxy’s negative bias grows in less-urbanized nonmetro areas (–8.3 min), indicating that the proxy understates travel time (overstates access) for remote block groups—plausibly because the straight-line-with-circuitry approach underestimates actual road detours in sparse rural road networks. The less-urbanized nonmetro misclassification rate of 19.4% ( $n = 31$ ) should be interpreted cautiously given the small stratum size. The dominant error direction is consistent with the overall pattern: the proxy tends to classify some truly-outside block groups as within access, meaning the true desert population is likely larger than reported.

**Table S18.** Proxy validation stratified by census division.

| Census division | $n$ | MAE (min) | RMSE (min) | Bias (min) | Misclass. rate |
| --- | --- | --- | --- | --- | --- |
| New England | 24 | 6.3 | 8.9 | –5.7 | 16.7% |
| Middle Atlantic | 67 | 4.9 | 5.9 | –3.5 | 9.0% |
| East North Central | 75 | 5.6 | 11.2 | –4.3 | 8.0% |
| West North Central | 32 | 3.6 | 4.4 | –0.9 | 3.1% |
| South Atlantic | 88 | 7.5 | 15.3 | –0.1 | 3.4% |
| East South Central | 25 | 8.0 | 12.1 | +2.0 | 8.0% |
| West South Central | 55 | 5.8 | 11.5 | +0.7 | 7.3% |
| Mountain | 34 | 13.7 | 24.4 | –4.5 | 2.9% |
| Pacific | 90 | 7.1 | 14.4 | –2.0 | 5.6% |

Notes: Mountain division has the highest MAE (13.7 min) and RMSE (24.4 min), consistent with the expectation that mountainous terrain inflates the gap between straight-line and network distances. Despite high error, the Mountain 30-minute misclassification rate is low (2.9%) because most Mountain block groups are clearly inside or outside the 30-minute threshold (few near-boundary cases). East South Central and West South Central are the only divisions where the proxy slightly overstates travel time (positive bias), with East South Central showing the larger overstatement.

**Table S19.** 30-minute threshold classification by terrain type.

| **Terrain group** | ***n*** | **Sensitivity** | **Specificity** | **Agreement** | **Misclass. rate** |
| --- | --- | --- | --- | --- | --- |
| Mountain division | 34 | 100.0% | 90.9% | 97.1% | 2.9% |
| Non-mountain divisions | 456 | 97.8% | 83.1% | 93.2% | 6.8% |
| Overall | 490 | 97.9% | 83.7% | 93.5% | 6.5% |

*Notes:* Sensitivity = true positive rate (correctly classified as within 30 min); specificity = true negative rate (correctly classified as outside 30 min). High sensitivity (97.9%) means the proxy rarely misses block groups that are truly within 30 minutes. The lower specificity (83.7%) means 16.3% of truly-outside block groups are misclassified as within access; this is the dominant error direction, consistent with the negative mean bias (proxy understates travel time). The inequality findings are conservative in the sense that the true desert population is likely larger than measured.

## Notes

### Competing Interest Statement

The authors have declared no competing interest.

### Funding Statement

This study did not receive any funding.

